# Prediction of CRT Response Using a Lead Placement Score Derived from 4DCT

**DOI:** 10.1101/2022.03.23.22272846

**Authors:** Ashish Manohar, Gabrielle M. Colvert, James Yang, Zhennong Chen, Maria J. Ledesma-Carbayo, Mads Brix Kronborg, Anders Sommer, Bjarne L. Nørgaard, Jens Cosedis Nielsen, Elliot R. McVeigh

**Author notes:** **Corresponding Author:** Elliot R. McVeigh, 9500 Gilman Dr, Mail Code: 0412, La Jolla, CA 92093.

## Abstract

**Background:** Cardiac resynchronization therapy (CRT) is an effective treatment for patients with heart failure; however, 30% of patients do not respond to the treatment. We sought to derive patient-specific left-ventricle (LV) maps of lead placement scores (LPS) that highlight target pacing lead sites for achieving a higher probability of CRT response.

**Methods:** Eighty-two subjects recruited for the ImagingCRT trial were retrospectively analyzed. All 82 subjects had two contrast-enhanced full-cardiac cycle 4DCT scans: a baseline and a 6-month follow-up scan. CRT response was defined as a reduction in CT-derived end-systolic volume ≥15%. Eight LV features derived from the baseline scans were used to train a support vector machine (SVM) via a bagging approach. An LPS map over the LV was created for each subject as a linear combination of the SVM feature weights and the subject’s own feature vector. Performance for distinguishing responders was performed on the original 82 subjects.

**Results:** Fifty-two (63%) subjects were responders. Subjects with an LPS≤Q_1_ (lower-quartile) had a posttest probability of responding of 14% (3/21), while subjects with an LPS≥ Q_3_ (upper-quartile) had a posttest probability of responding of 90% (19/21). Subjects with Q_1_<LPS<Q_3_ had a posttest probability of responding that was essentially unchanged from the pretest probability (75% vs 63%, p=0.2). An LPS threshold that maximized the geometric mean of true-negative and true-positive rates identified 26/30 of the non-responders. The AUC of the ROC curve for identifying responders with an LPS threshold was 87%.

**Conclusions:** An LPS map was defined using 4DCT-derived features of LV mechanics. The LPS correlated with CRT response, reclassifying 25% of the subjects into low-probability of response, 25% into high-probability of response, and 50% unchanged. These encouraging results highlight the potential utility of 4DCT in guiding patient selection for CRT. The present findings need verification in larger independent data sets and prospective trials.

**Clinical Perspective:** Cardiac resynchronization therapy (CRT) is a proven treatment for patients with heart failure and dyssynchrony; however, approximately 30% of patients do not respond to the treatment. Additionally, the relatively high non-responder rate poses difficulties for the optimal utilization of medical resources; thus, more accurate patient stratification for CRT remains an unmet need. Despite significant efforts focused on using imaging to guide CRT, the results thus far have been ambiguous. Poor reproducibility of echocardiography coupled with the complexity of cardiac magnetic resonance have likely contributed to the poor overall adoption of these methods for pre-CRT assessment. In this work, we describe a metric called the lead placement score (LPS) that combines multiple 4DCT-derived features of left-ventricular (LV) mechanics into a single number for each possible pacing lead location on the LV; the features included in the LPS map have previously been shown to correlate with CRT response. Using a machine learning classifier, a model was constructed with these features and then used to derive the LPS map for each individual subject. The LPS was found to correlate with the probability of a subject responding to CRT. 4DCT is widely available and provides high-resolution images of the full cardiac cycle. Additionally, recent technological advancements have also dramatically reduced the radiation dose from 4DCT scans. The advantages of 4DCT coupled with the promising results reported in this study, highlight the potential utility of 4DCT in the planning of CRT.

## 1. Introduction

Multiple trials have proven that cardiac resynchronization therapy (CRT) can provide significant benefit for patients with intraventricular dyssynchrony and heart failure [1]. Current guidelines for CRT patient selection include New York Heart Association (NYHA) functional classes II-IV, echocardiography-derived left ventricular ejection fractions (LVEF) ≤ 35%, and QRS durations >120 ms [2]. Despite its proven benefit, approximately 30% of patients selected for CRT do not respond to the treatment [3]. This non-responder rate continues to pose considerable challenges for the optimal utilization of medical resources [4]; thus, necessitating accurate patient stratification.

Significant effort has been focused on reducing the non-responder rate through the use of imaging, for the most part with echocardiography [5]. Two-dimensional radial strain from speckle-tracking echocardiography has been previously used to identify optimal left ventricle (LV) lead placement sites [6]. Additionally, a number of studies have explored the role of tissue doppler imaging in patient selection for CRT [7]. However, poor reproducibility of echocardiography measurements due to inter-and intra-observer variability coupled with inter-vendor differences have led to disappointing results and hindered its routine clinical use [8].

Similarly, cardiac magnetic resonance (CMR) imaging has also been used to guide CRT [9]. Cine MRI [10], tagged MRI [11], and cine DENSE [12] have shown great promise in mapping regional strain and mechanical activation times of the LV myocardium. Also, late gadolinium enhancement imaging is effective at identifying regions of scar tissue that need to be avoided for lead placement [13]. However, 28% of patients in consideration for CRT already have existing right ventricular (RV) pacing systems in place [2], making CMR not widely available to this cohort. Additionally, tagged MRI and cine DENSE images are not simple to acquire and analyze, requiring skilled technicians and image analysis personnel. No imaging modality is currently recommended for CRT planning and management.

Recent studies have explored the use of 4DCT to guide CRT. Truong et al. used dual-source CT to derive LV dyssynchrony indices that predicted 2-year major adverse cardiac events (MACE). They also reported that leads placed on sites with maximal wall thickness correlated with less MACE [14]. Rinaldi and colleagues utilized 4DCT-derived assessment of LV dyssynchrony and myocardial scar to target LV lead placement. They showed that patients with leads implanted in segments targeted from CT had higher clinical response rates [15] and superior acute hemodynamic responses [16] than those with leads implanted in the non-target segments. Fyenbo et al. used CT to identify regions of myocardial scar and to compute scar burden. They found that high scar burden and proximity of scar to the LV pacing site were correlated with echocardiographic non-response [17].

We hoped to improve on the insights from these previous studies by using additional features of LV mechanics that have previously been shown to correlate with CRT response[1] in a larger number of subjects. Thus, the purpose of this study was to use a combination of 4DCT-derived regional and global features of LV mechanics to define patient-specific maps of lead placement scores (LPS) that are correlated with CRT response.

## 2. Methods

### 2.1 Subject Population and CT Imaging

Subjects recruited for the ImagingCRT [18] randomized controlled trial were retrospectively used for this study. The trial enrolled a total of 182 subjects; however, only 147 subjects had contrast enhanced 4DCT scans both before and after CRT implantation. The complete study protocol for the trial is described in detail by Sommer et al. [18], [19]. Briefly, all 147 subjects were > 40 years, belonged to NYHA functional classes II-IV, had echocardiography derived LVEF ≤ 35%, and had QRS widths ≥ 120 ms with a left bundle branch block (LBBB) or > 180 ms with an RV pacing configuration, respectively. The trial was conducted at the Department of Cardiology, Aarhus University Hospital, Skejby, Denmark and was approved by the Central Denmark regional committee on health research ethics as well as the Danish Data Protection Agency. All trial participants gave informed written consent, and the trial was registered on Clinicaltrials.gov (NCT01323686).

The prescribed cardiac 4DCT imaging protocol has previously been described in detail [18]. Each subject had two contrast-enhanced full cardiac cycle and full LV volume retrospective ECG-gated 4DCT scans: the first scan was on the day prior to CRT implantation and the second scan was 6-months after implantation; we refer to these scans hereafter as the *baseline* and the *follow-up* scans, respectively. All scans were acquired using a dual-source CT scanner (Somatom Definition Flash, Siemens Healthcare, Erlangen, Germany) with a multi-slice z-axis detector of 38.4 mm, a gantry rotation time of 280 ms, pitch factors in the range of [0.17, 0.3], x-ray tube voltages of 80-120 kVp, and adaptive tube currents with a reference of 370 mAs. In these retrospective scans, an ECG controlled tube current modulation was applied with reduction of the current to 20% and reference mA applied only between 60-70% of the R-R interval. Images were reconstructed every 5% of the R-R interval, from 0% to 95%, using an iterative reconstruction algorithm (SAFIRE I26f-4, Siemens Healthcare, Erlangen, Germany). The reconstructed images had in-plane pixel spacings in the range of [0.6, 1] mm and slice thicknesses of 0.7 mm.

Subjects were excluded from this study if either their baseline or follow-up scans had one or more of the following imaging artifacts, preventing precise measurements of LV mechanics:

1. Severe helical step artifact
2. Insufficient LV chamber-myocardium contrast for blood volume segmentation
3. Severe metal artifacts from the pacemaker/defibrillator leads preventing segmentation of the LV blood volume

### 2.2 Image Segmentation and Mesh Extraction

The LV blood pool was segmented from the reconstructed CT images in a manner similar to that described by Colvert et al. [20] and Manohar et al. [21]. Briefly, the blood pool was segmented in ITK-SNAP v3.8.0 [22] software using the active contour region growing module. Two planes delineating the mitral valve and the left ventricular outflow tract (LVOT) were defined as previously described [20]. The process was repeated for all 20 time frames in each of the baseline and the follow-up 4DCT datasets of each subject. Additionally, the positions of the right and the left lead tips were marked in the end diastolic image of the follow-up scans; these positions were used to project the corresponding right and left pacing sites onto the LV endocardial model, respectively. Figure 1 illustrates the segmentation process.

**Fig. 1.**
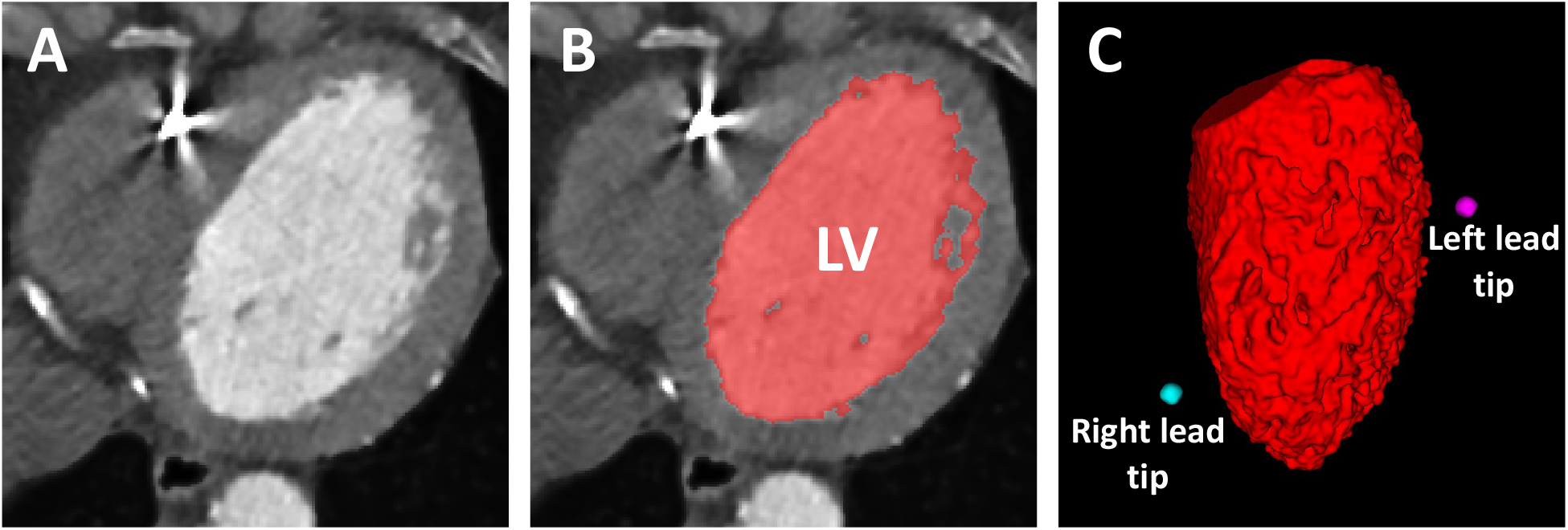
LV blood-volume segmentation. (**A**) Axial contrast-enhanced cardiac CT image. (**B**) LV blood-volume segmentation overlaid in red. (**C**) 3D rendering of the LV blood-volume segmentation with the locations of the right (cyan) and the left (magenta) lead tips. The anterior wall is in view with the septum on the left and the lateral wall on the right. LV: left ventricle.

Meshes defining the endocardium were extracted using the *isosurface* MATLAB routine (MathWorks Inc., Natick, MA; v9.8 – R2020a). Vertices and faces belonging to the defined mitral valve and LVOT planes were removed, leaving a 2D surface delineating only the LV endocardium. The meshes were extracted at an isotropic resolution of 2 mm in x, y, and z directions for calculating local LV function, which was consistent with previously published analyses [20], [21], [23].

### 2.3 Global and Regional Features of LV Mechanics

Eight global and regional features of LV mechanics were derived from the baseline 4DCT scans for each subject. The global LV features used were:

1. End diastolic volume (EDV)
2. End systolic volume (ESV)
3. Circumferential uniformity ratio estimate using singular value decomposition (CURE-SVD),
4. LV sphericity index (LVSI)

and the regional LV features used were:

1. Peak regional shortening (PRS_CT_)
2. Time to peak regional shortening (TPRS_CT_)
3. Maximum pre-stretch of regional shortening (MSRS_CT_)
4. Time to onset of shortening (TOS)

The choice of these 8 features was driven by previous CRT studies using CMR and CT imaging [1], [10]. Detailed information on the computation of the above 8 features can be found in the Supplemental Material. Figure S1 describes the estimation of the 4 regional LV features.

### 2.4 Spatial Sampling of the LV Endocardium

The LV endocardium for each subject was divided into 90 spatial segments: 18 circumferential segments (one every 20°) for each slice, and 5 slices defined from apex to base along the long axis of the LV [20]. This 90 segment model yields a higher spatial sampling than the traditional AHA 17 segment model, permitting wall function analysis that captures the high resolution features of mechanical function that are obvious in the 4DCT data. The location of the right and the left lead tips were mapped onto the LV endocardial segments closest to the lead locations.

### 2.5 Definition of CRT Response

Subjects were considered responders if their CT-derived ESV decreased by ≥ 15% 6-months post CRT implantation [8]. The ESV of the subjects were computed from the 4DCT exams as described in the Supplemental Material and the change in ESV 6-months post implantation was calculated between the baseline and the follow-up 4DCT scans.

### 2.6 Lead Placement Score

The LPS is a scalar parameter that is obtained by combining several features derived from the baseline 4DCT scan of a subject under consideration for CRT. Our goal was to derive an LPS that is correlated with CRT response. An individual LPS can be calculated for each of the 90 segments of the LV endocardium, yielding a high-resolution patient-specific LPS map that highlights possible target pacing lead sites for achieving a higher probability of CRT response. Since all subjects in this study had follow-up 4DCT scans, the locations of the right and the left lead tips were precisely known; thus, to match the response observed in these subjects, the LPS value in our analysis was obtained from the right and left lead tip locations projected onto the baseline 4DCT scans.

The LPS values were computed using a model obtained by training a support vector machine (SVM) [24]. The following features derived from the baseline scans of the subjects were used in model training: the global LV features of 1) EDV, 2) ESV, 3) CURE-SVD, and 4) LVSI, and the regional LV features of 5) PRS_CT_, 6) TPRS_CT_, 7) MSRS_CT_, and 8) TOS. For each of the regional LV features, the values at the two spatial segments in the baseline scans that corresponded to the right and the left lead tip locations in the follow-up scans were used to calculate a single LPS value for each subject. For example, TOS-right and TOS-left represented the TOS of the LV segments that spatially corresponded to the segments of the right and the left lead tips in the follow-up scans. Thus, a total of 12 features were used to train the SVM model (4 global + 4×2 regional). Detailed information on the SVM training and the LPS calculation can be found in the Supplemental Material.

## 3. Results

### 3.1 Study Population

Out of the 147 subjects that had 4DCT scans acquired at both baseline and at 6-months follow-up, 82 subjects had images that were of sufficient quality to be usable for this study. Scans of subjects were excluded because of either helical step artifacts (37 subjects, 25%), insufficient LV blood pool-myocardium contrast (19 subjects, 13%), or severe metallic lead artifacts (9 subjects, 6%). Figure 2 shows a flow diagram of the subject selection process used in this study. The mean radiation dose across the 82 subjects was 4.4 ± 2.6 mSv (median: 4.1 mSv; IQR: 2 mSv). The baseline characteristics are provided in Table 1.

**Fig. 2.**
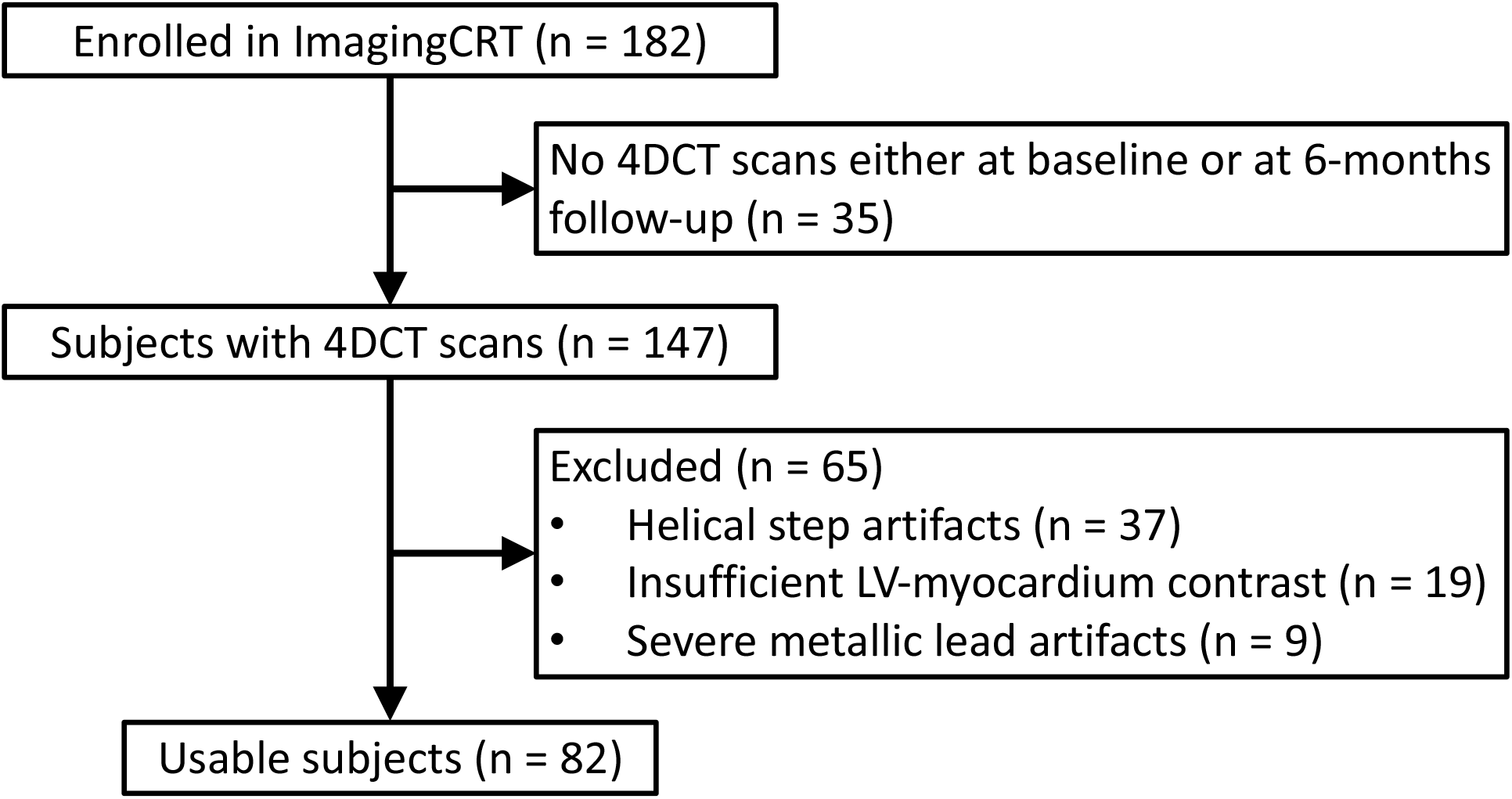
Flow diagram of the subject selection process.

**Table 1.**
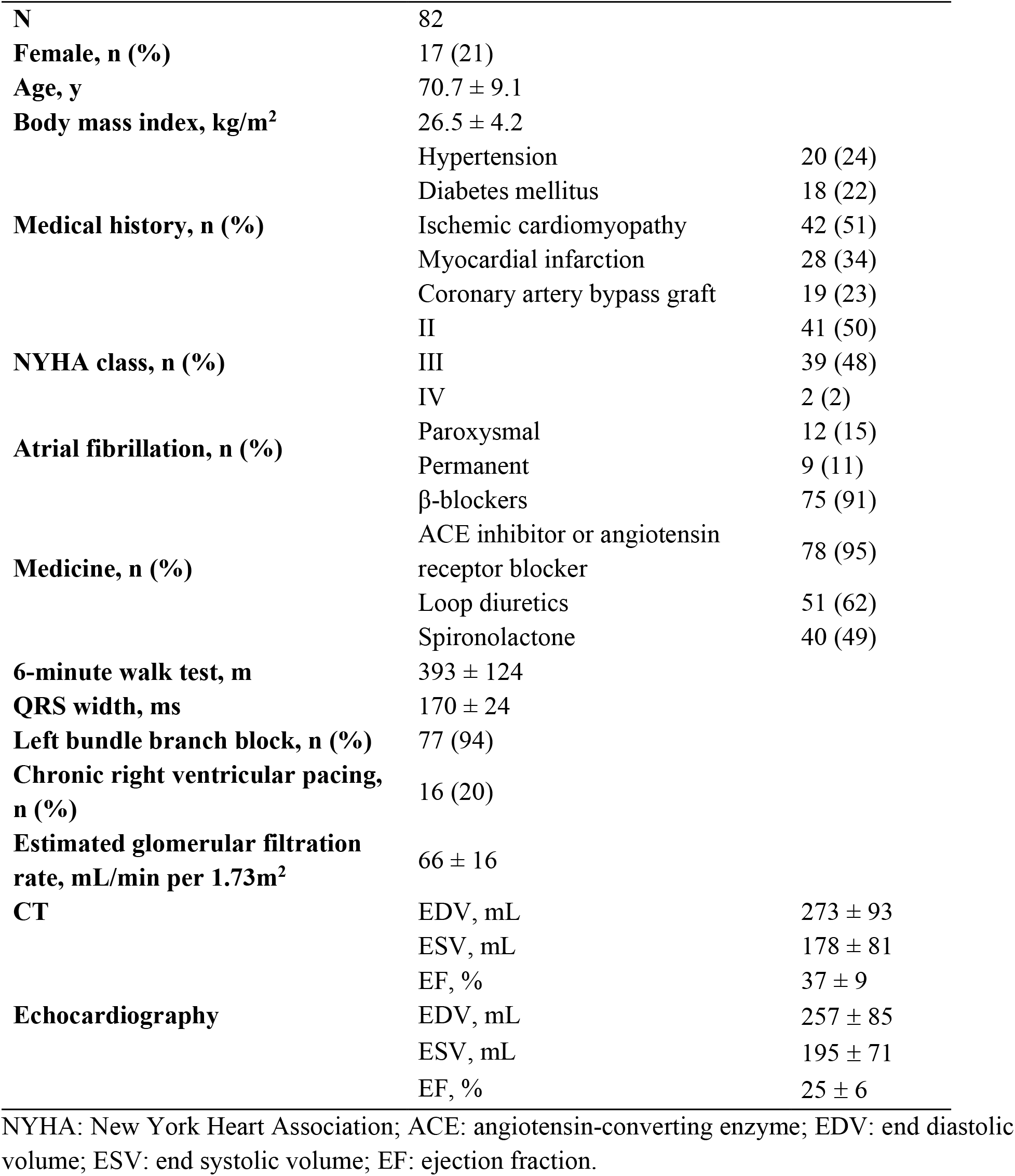
Baseline characteristics of the 82 subjects.

The CT-derived and the echocardiography-derived LVEFs at baseline were statistically different (37 ± 9% vs 25 ± 6%, p < 0.001) and poorly correlated (r = 0.46, p < 0.001).

### 3.2 Lead Placement Score and Response Prediction

Out of the 82 subjects, 52 (63%) had an ESV decrease of ≥ 15% at 6-months follow-up and were considered responders. The average weights of the 12 features that were used in training the SVM model are shown in Table 2.

**Table 2.**
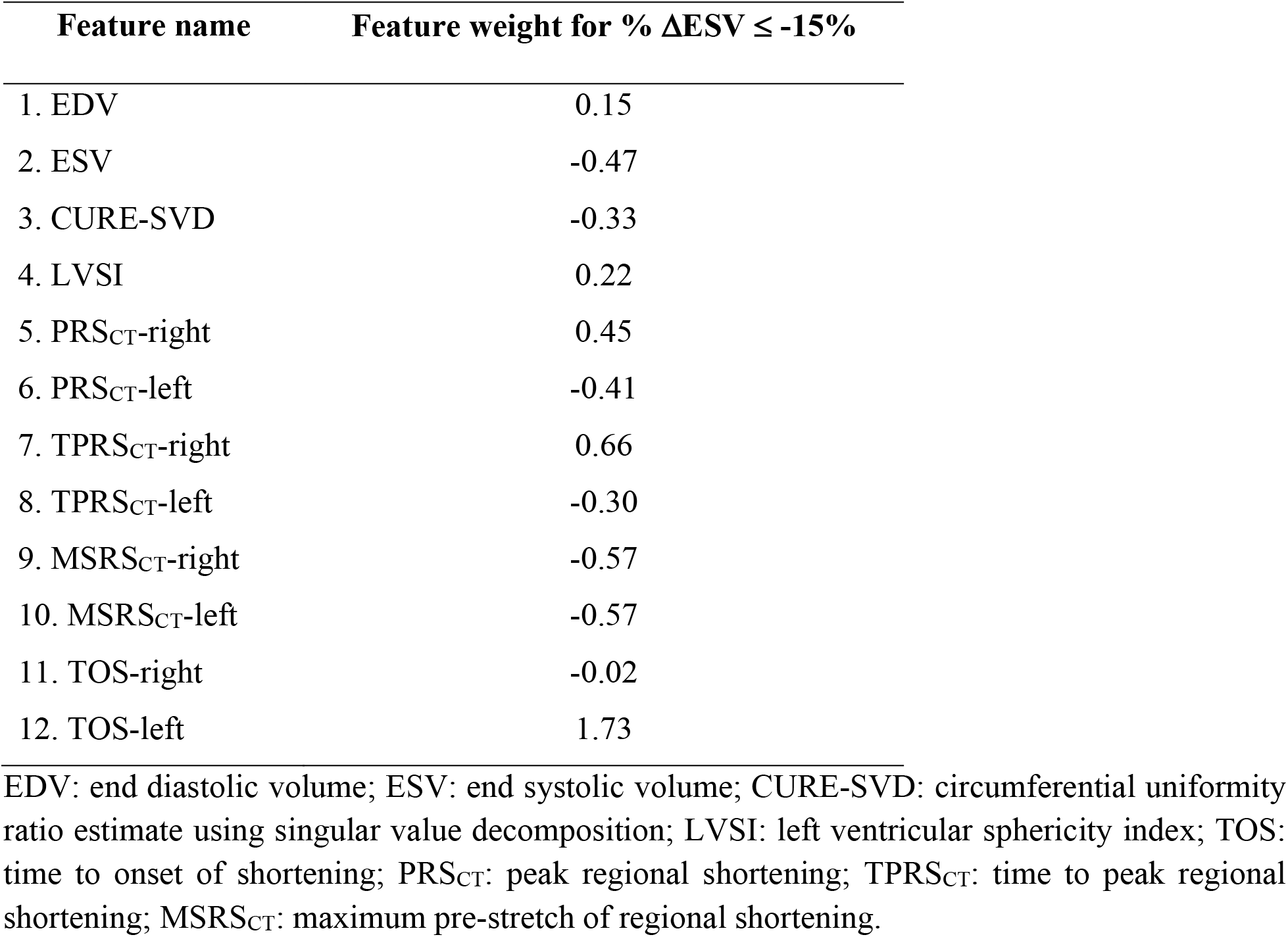
Feature weights of the SVM model with the responder criterion set at % ΔESV ≤ -15%.

| Feature name | Feature weight for $\% \Delta\text{ESV} \leq -15\%$ |
| --- | --- |
| 1. EDV | 0.15 |
| 2. ESV | -0.47 |
| 3. CURE-SVD | -0.33 |
| 4. LVSI | 0.22 |
| 5. PRS<sub>CT</sub>-right | 0.45 |
| 6. PRS<sub>CT</sub>-left | -0.41 |
| 7. TPRS<sub>CT</sub>-right | 0.66 |
| 8. TPRS<sub>CT</sub>-left | -0.30 |
| 9. MSRS<sub>CT</sub>-right | -0.57 |
| 10. MSRS<sub>CT</sub>-left | -0.57 |
| 11. TOS-right | -0.02 |
| 12. TOS-left | 1.73 |

EDV: end diastolic volume; ESV: end systolic volume; CURE-SVD: circumferential uniformity ratio estimate using singular value decomposition; LVSI: left ventricular sphericity index; TOS: time to onset of shortening; PRS<sub>CT</sub>: peak regional shortening; TPRS<sub>CT</sub>: time to peak regional shortening; MSRS<sub>CT</sub>: maximum pre-stretch of regional shortening.

Figure 3 shows the relationship between the LPS and the relative change in ESV between the baseline and the follow-up scans. An LPS threshold of -0.2 maximized the geometric mean (g-mean) value for response prediction; among the subjects above this threshold, the non-responder rate was 9% (4/(4+41)), down from the original non-responder rate of 37% (30/82), and the sensitivity and the specificity at this threshold were 79% and 87%, respectively.

**Fig. 3.**
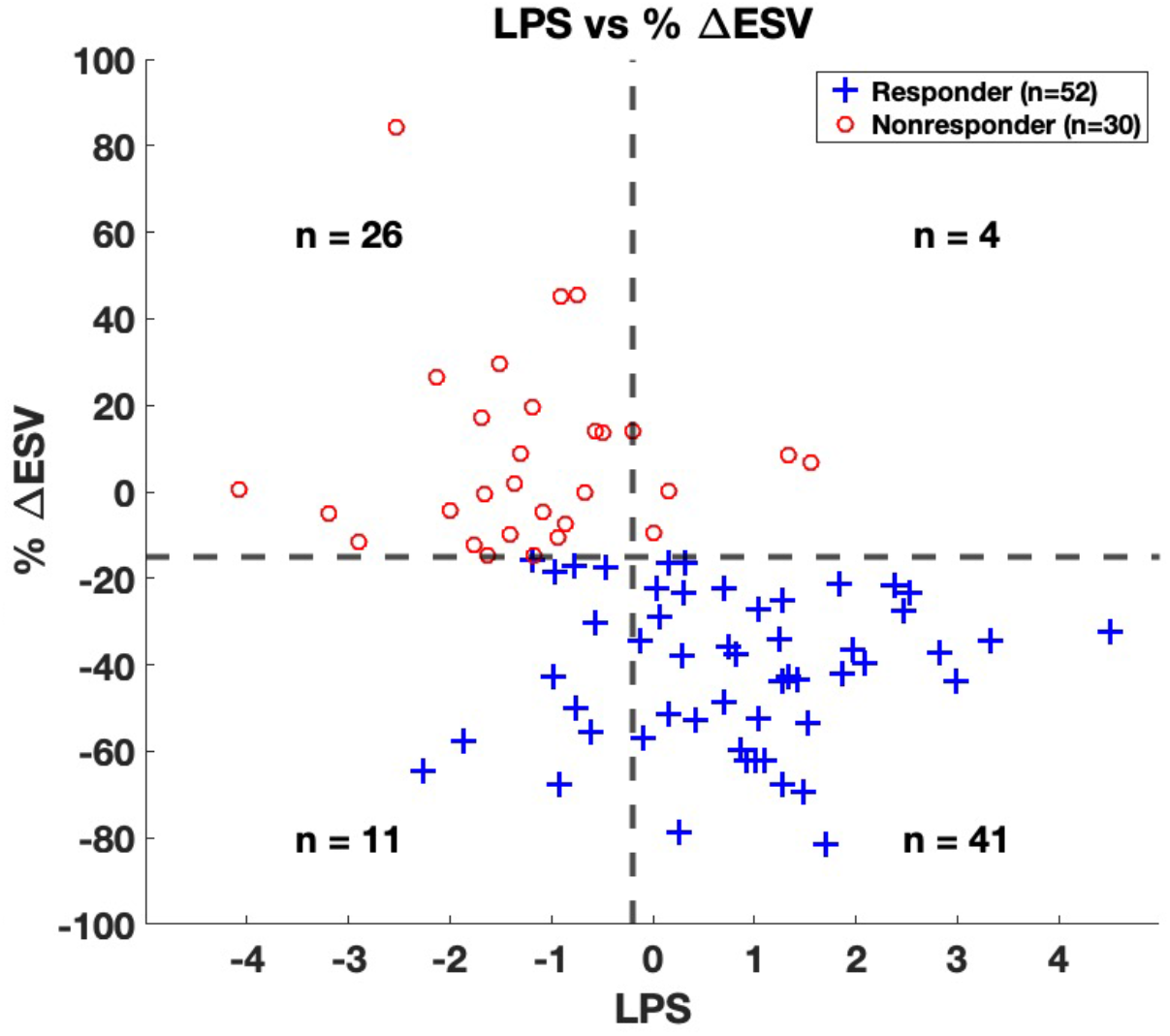
Relationship between change in ESV at 6-months follow-up and lead placement score (LPS). Distribution of LPS values for the lead locations in each subject as a function of the relative change in ESV between baseline and follow-up scans. The vertical dashed line corresponds to the LPS threshold (−0.2) that maximized the g-mean value for response prediction. The horizontal dashed line corresponds to the response definition of a relative reduction in ESV of 15%. ‘+’ represents a true responder and ‘o’ represents a true non-responder. LPS: lead placement score; ESV: end systolic volume.

Figures 4A shows the histogram of the LPS values for all 82 subjects. The LPS of the true non-responders are shown in red while those of the true responders are shown in blue. Figure 4B shows the receiver operating characteristic (ROC) curve of the prediction model. The area under the curve (AUC) was 87% (95% confidence interval: 77 – 94%).

**Fig. 4.**
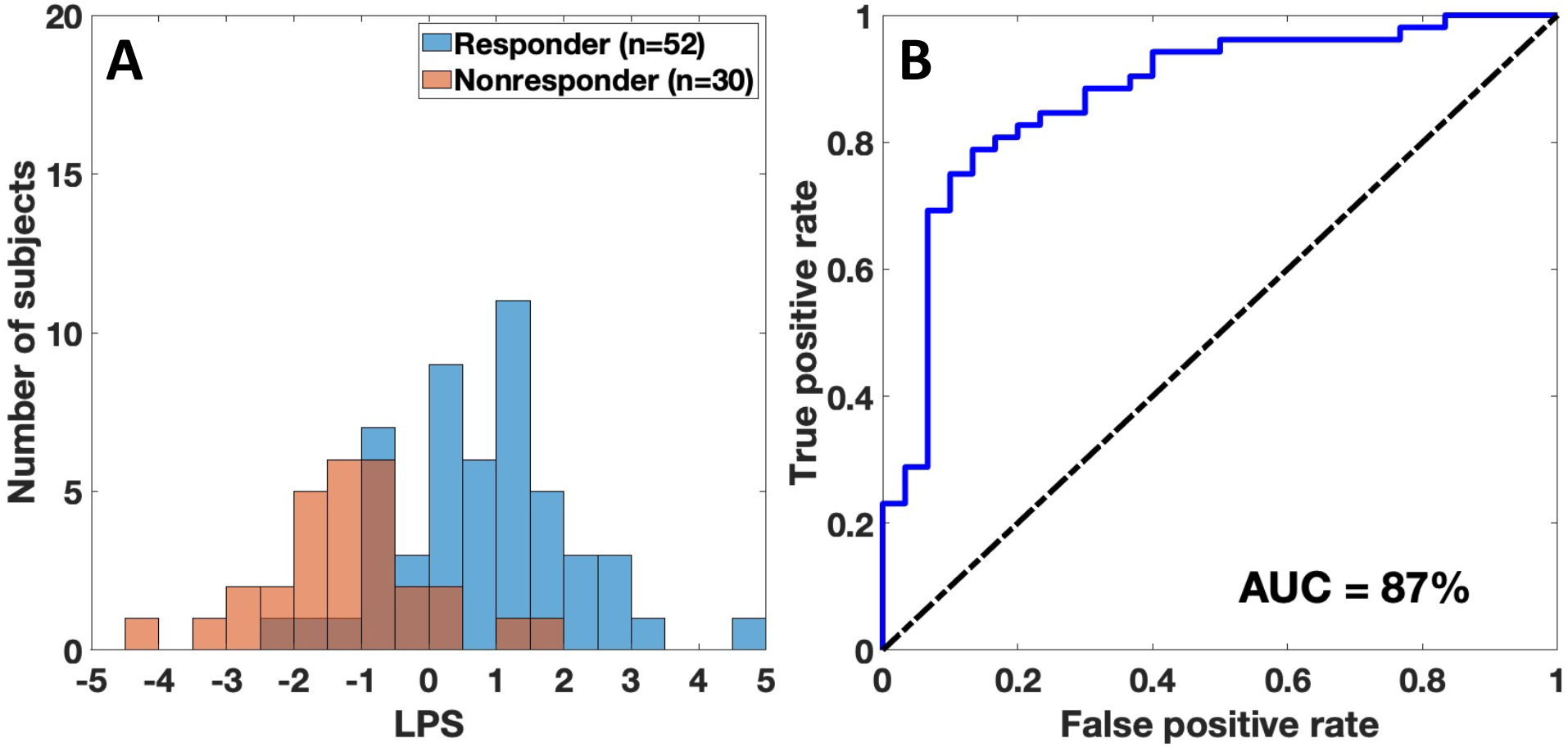
(**A**)Histogram of LPS for all 82 subjects. The non-responders are shown in red, and the responders are shown in blue. (**B**) ROC curve of the trained SVM model to predict responders as a function of changing LPS threshold. LPS: lead placement scores; ROC: receiver operating characteristic; AUC: area under curve; SVM: support vector machine.

There is a very practical way to stratify the subjects using the LPS. We can break the subjects into 3 groups with respect to their posttest probabilities: lower probability of response, unchanged probability, and higher probability of response. Without considering the 4DCT data, the pretest probability of responding to CRT using the ≥15% reduction in ESV definition was 52/82 (63%). For the same response definition, by incorporating the 4DCT data, subjects with an LPS in the lowest quartile (LPS ≤ Q_1_ = -1) had a posttest probability of responding of 14% (3/21). Similarly, subjects with an LPS in the highest quartile (LPS ≥ Q_3_ = 1.3) had a posttest probability of responding of 90% (19/21), and for those subjects with an LPS within the interquartile range (IQR; Q_1_ < LPS < Q_3_), the posttest probability remained essentially unchanged from the pretest probability (75% vs 63%; p = 0.2).

The above analysis was performed on subjects that had already undergone CRT; hence, the precise locations of the left and the right lead tips were known. For our analysis of predicting response in these subjects, we used features of LV mechanics that were estimated from the baseline 4DCT scans at the two endocardial segments corresponding to the pacing lead tip locations. However, in general, the analysis pipeline developed can be applied to generate high resolution LV maps of LPS, aiding in the selection of target lead placement sites for achieving a higher probability of response. Fixing the location of the right lead, LPS maps were generated for the LV free wall (anterior wall to inferior wall).

Figure 5 shows four example subjects that highlight the significance of the LPS in identifying target lead placement sites: (a) a responder (%ΔESV =-43) with uniformly high LPS values across the entire LV free wall and the left lead placed in this region of high LPS, (b) a responder (%ΔESV = -68) with a localized region of high LPS values on the basal inferolateral wall and the left lead placed in this region of high LPS, (c) a non-responder (%ΔESV = -13) with globally low LPS values and thus the left lead placed in a region of low LPS, and (d) a non-responder (%ΔESV = +14) with a localized region of high LPS values on the basal inferolateral wall, but the left lead not placed in this region of high LPS. For each subject, polar maps of the 4 regional features of LV mechanics (TOS, PRS_CT_, TPRS_CT_, and MSRS_CT_) that were used in the derivation of the LPS are shown. Additionally, the location of the right and the left lead tips are shown by the blue and red highlighted segments, respectively. Also shown are polar maps of the LPS as well as 3D renderings of the LV lateral wall with the LPS values mapped onto the endocardial surface.

**Fig. 5.**
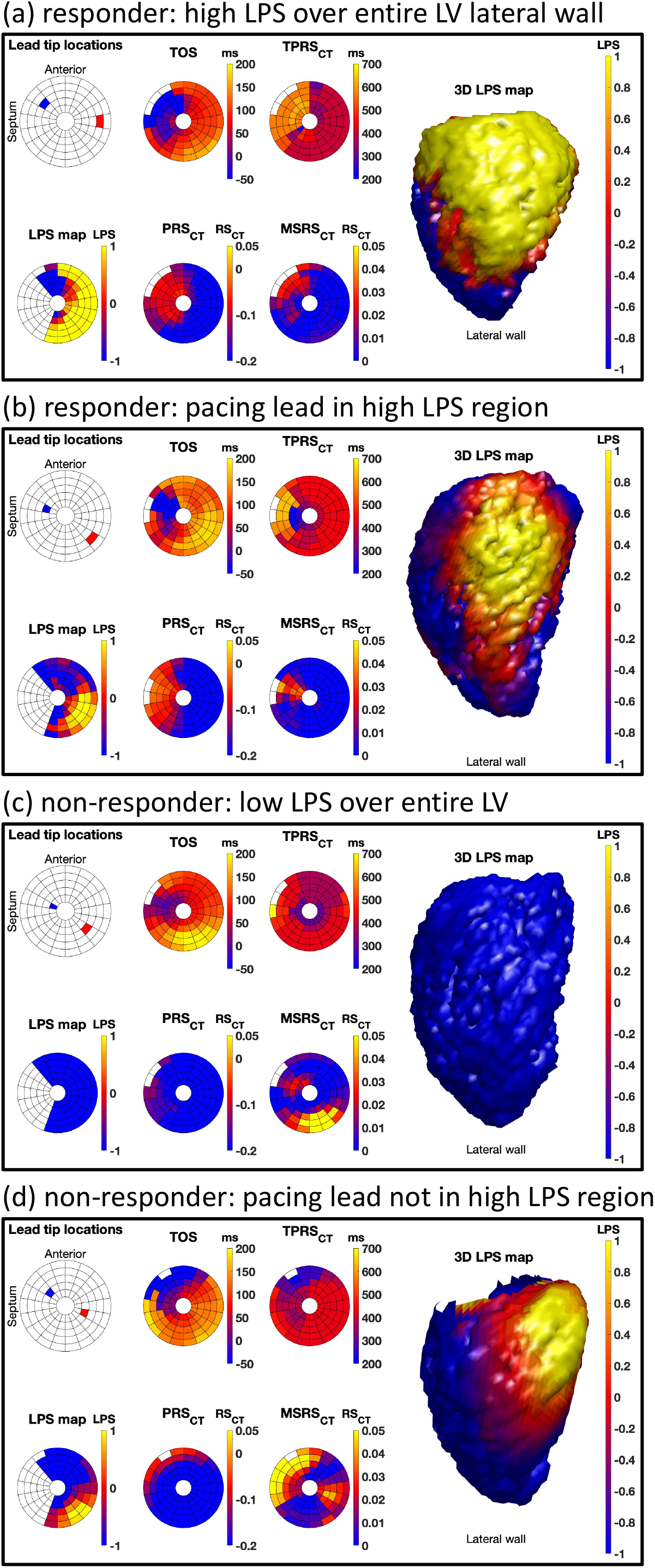
Lead placement score (LPS) maps of CRT responders and non-responders. (**a-d**) Four example subjects with the following information shown for each subject **Lead tip locations**: blue = right lead, red = left lead **TOS**: time to onset of shortening, in ms; **PRS**_**CT**_: peak regional shortening; **TPRS**_**CT**_: time to peak regional shortening, in ms; **MSRS**_**CT**_: maximum pre-stretch of regional shortening; **LPS map**: lead placement score map as a polar map; **3D LPS map**: 3D rendering of the LV lateral wall with LPS values mapped onto the endocardial surface. **(a)** a responder (%ΔESV = -43) with uniformly high LPS values across the entire LV free wall and the left lead placed in this region of high LPS **(b)** a responder (%ΔESV = -68) with a localized region of high LPS values on the basal inferolateral wall and the left lead placed in this region of high LPS **(c)** a non-responder (%ΔESV = - 13) with globally low LPS values and thus the left lead placed in a region of low LPS **(d)** a non-responder (%ΔESV = +14) with a localized region of high LPS values on the basal inferolateral wall, but the left lead not placed in this region of high LPS

## 4. Discussion

The primary objective of this study was to use known quantitative features of LV mechanics derived from baseline 4DCT scans as features in a *lead placement score* that correlated with CRT response. The main findings reported demonstrate the potential utility of 4DCT in guiding patient selection for CRT; in at least 42 of the 82 subjects used in the study, the LPS could have a significant impact on the decision to proceed with CRT. The study also highlights the patient-specific variation in the distribution of LPS values across the LV; some subjects had large areas with high LPS values, providing multiple LV lead placement target sites for achieving a higher probability of CRT response. Other subjects had small areas with high LPS values; knowing this information may be beneficial in designing CRT delivery that is most effective. Lastly, some subjects had uniformly low LPS values across the LV; CRT may not be the optimal treatment for these subjects, especially since CRT is not without risk. Thus, the patient-specific LPS maps could aid in the optimal planning and management of patients under consideration for CRT.

4DCT scans of 147 subjects recruited for the ImagingCRT trial were retrospectively analyzed in this study. While the temporal resolution of the CT images was high because of the “dual-source” technology of the particular CT scanner used to acquire the 4DCT images, due to the limited z-axis coverage, imaging the entire superior-inferior extent of the heart was performed in helical mode. Unfortunately, 37 of the 147 subjects (25%) had severe step artifacts due to beat-to-beat irregularities, rendering the images not usable for dyssynchrony analysis. Despite the excellent temporal resolution of the dual-source scanner (66 ms per frame), wider detector scanners (256 or 320 detector-rows) with full heart coverage from a single table position may be better suited for this application, especially with the recent innovations in motion estimation and motion compensation technology [25], [26].

Despite 30 years of clinical development, no single definition of CRT response has been universally accepted [4]; however, ≥15% reduction in LV ESV is the most commonly used [2], [8] and can be measured with great precision using 4DCT. Previous studies using CT to guide CRT have used echocardiography derived LV ESV to determine response [16], [27]; however, the reproducibility of the echocardiography measures is low causing uncertainty in the results. The image-based response definition used in this study was derived from the LV blood volume segmentations of the baseline and the follow-up 4DCT images, which are *highly* reproducible [28] and are free from any assumptions about LV geometry or manual contouring. For these reasons, we are confident in the precision of the 4DCT derived measurement of change in ESV as mechanical response. Additionally, the mean baseline CT-derived LVEF was higher than the echocardiography-derived LVEF due to the simple “blood pool voxel” counting used to compute LVEF from CT; this bias was expected.

To the best of our knowledge, this is the first study that combined multiple features derived from a baseline 4DCT scan into a single scalar value (the “lead placement score”) that is correlated with CRT response. In addition, this paper differs from previous work using 4DCT to guide CRT in that we computed traditional dyssynchrony parameters (CURE, TOS) that have previously demonstrated a correlation with CRT response. The 4DCT scans contain an abundance of information; using a limited number of features derived from the scans for training, an SVM model was able to stratify subjects into LV mechanics responders and non-responders with high accuracy. An SVM was chosen as the desired classifier because: 1) it is less susceptible to overfitting [29] and 2) it has no local minima because it is defined by a convex optimization problem. Due to the limited number of subjects in this study, coupled with the class imbalance between the responders and the non-responders, it was not feasible to have an independent testing dataset. Thus, we implemented a bagging (bootstrap aggregation)[30] approach to overcome this limitation; bagging is an established method that improves stability and helps with overfitting [31].

The LPS was defined using a model that comprised 8 global and regional features of LV mechanics. TOS-left (time to onset of shortening at the left lead location) was the most prominent feature of the model, with its feature weight nearly three times that of the second most prominent feature. EDV, ESV, and LVSI are features that describe the size and shape of the LV. While a reduction in ESV is itself used as an established definition of CRT response, previous studies have investigated the relationship between CRT response and EDV and LVSI [32], [33]. Dyssynchrony features such as CURE [34], TOS [12], and TPRS_CT_ [35] have all been shown to correlate with CRT response; the results from this study are consistent with these previously published results. Peak shortening (PRS_CT_) and maximum pre-stretch (MSRS_CT_) are features that capture the function and viability of an LV myocardial region. Low values of PRS_CT_ are useful in identifying regions of scar/ischemic tissue, much like late gadolinium enhancement and reduced wall thickening; pacing at these sites is to be avoided as they are correlated with CRT non-response [1]. To the best of our knowledge, the influence of maximum pre-stretch (MSRS_CT_) on CRT response has not been previously reported; the results reported in this study reveal it to be a relatively important feature of the model used to stratify subjects for CRT. A large pre-stretch at either the location of the right or left lead likely detects a region of poor myocardial health which confers the observed reduction in probability of response.

The analysis pipeline adopted in this study was fully automated except for the selection of the mitral valve plane and the boundary plane between the LV chamber and the aortic outflow tract, which were performed manually using an algorithm derived from selected morphological features. The inter- and intra-user variability of this segmentation process has been previously shown to be very low, making RS_CT_ a highly reproducible metric for estimating regional endocardial function [20]. Additionally, with the advancements made in the field of deep learning cardiac image segmentation [36], [37], the entire segmentation component of the analysis pipeline will likely be fully automated in the near future.

The role of imaging in guiding CRT has been uncertain. A potential reason for this is the high variability of measurements obtained from echocardiography [8]. Modern 4DCT imaging systems can acquire high resolution images of the entire heart across the full cardiac cycle very rapidly, and in the case of wide detector systems, within a single heartbeat. Therefore, these systems do not suffer from step artifacts; thus, enabling artifact-free imaging of patients with arrhythmias. Another benefit of 4DCT is its ability to image patients with implanted metallic medical devices; nearly 28% of patients in consideration for CRT have existing RV pacing systems in place [2]. Additionally, using the dynamic mA feature of the scanner, the subjects used in this study had low CT-based radiation doses (median: 4.1 mSv; IQR: 2 mSv), which is comparable to the dose received from natural sources of radiation annually [38]. The dose from CT is continuously being reduced as CT technology advances; from the 2007 and the 2017 dose surveys, the dose from coronary CT angiography was reduced by 78% (885 mGy*cm vs 195 mGy*cm, p < 0.001) [39]. The limits of achievable dose reduction are further being pushed by technological advancements such as iterative reconstruction, photon-counting detectors, and the use of deep learning [40].

### 4.1 Limitations

This study uses 4DCT scans of 82 subjects acquired at both baseline and at 6-months follow-up after CRT implantation. This is a unique dataset of 4DCT scans where a highly precise direct comparison of LV mechanical states can be made pre- and post-CRT. While the models derived in this study show great promise for using 4DCT in guiding patient selection for CRT, analysis of independent data and prospective trials will be required to ensure generalizability. CRT is now an accepted therapy for dyssynchronous heart failure; therefore, larger datasets are available for testing our existing model.

All 4DCT images used in this study were acquired and reconstructed with the same CT imaging system (Somatom Definition Flash, Siemens Healthcare, Erlangen, Germany); this was tremendously advantageous in facilitating a direct comparison of LPS values derived from 4DCT scans of similar image characteristics. Future studies will need to explore the effect of vendor specific differences and total radiation dose on the computed LPS values.

## 5. Conclusions

A lead placement score map was developed using features of LV mechanics that were derived from 4DCT images acquired in 82 subjects prior to CRT implantation. The LPS value at the lead locations correlated with subject response to CRT; it effectively stratified 1/4 of the subjects into low probability (14%) of response and 1/4 into high probability (90%) of response, and the remaining 1/2 had a probability of response that was unchanged from the pretest probability (75% vs 63%, p = 0.2). Additionally, an LPS threshold that maximized the geometric mean of true negative and true positive rates categorized subjects as responders and non-responders with high sensitivity (79%) and specificity (87%), with an area under the ROC curve of 87% (95% CI: 77 - 94%). These encouraging results highlight the potential utility of 4DCT in planning CRT.

## Data Availability

All data produced in the present study are available upon reasonable request to the authors

## Non-standard Abbreviations and Acronyms

CRT: cardiac resynchronization therapy
NYHA: New York Heart Association
LVEF: left ventricular ejection fraction
LV: left ventricle
CMR: cardiac magnetic resonance
RV: right ventricle
MACE: major adverse cardiac events
LPS: lead placement score
LBBB: left bundle branch block
LVOT: left ventricular outflow tract
EDV: end diastolic volume
ESV: end systolic volume
CURE: circumferential uniformity ratio estimate
SVD: singular value decomposition
CURE-SVD: circumferential uniformity ratio estimate using singular value decomposition
LVSI: left ventricular sphericity index
RS_CT_: regional shortening
PRS_CT_: peak regional shortening
TPRS_CT_: time to peak regional shortening
MSRS_CT_: maximum stretch of regional shortening
TOS: time to onset of shortening
SVM: support vector machine
IQR: interquartile range
ROC: receiver operating characteristic
AUC: area under curve
CI: confidence interval

## Supplemental Material

Supplemental methods

Figure S1

References 41-48

## References

[1] F. W. Prinzen, K. Vernooy, and A. Auricchio, “Cardiac Resynchronization Therapy,” Circulation, vol. 128, no. 22, pp. 2407–2418, Nov. 2013, doi: 10.1161/CIRCULATIONAHA.112.000112.

[2] J.-C. Daubert et al., “2012 EHRA/HRS expert consensus statement on cardiac resynchronization therapy in heart failure: implant and follow-up recommendations and management: A registered branch of the European Society of Cardiology (ESC), and the Heart Rhythm Society; and in col,” Europace, vol. 14, no. 9, pp. 1236–1286, Sep. 2012, doi: 10.1093/europace/eus222.

[3] K. Vernooy, C. J. M. van Deursen, M. Strik, and F. W. Prinzen, “Strategies to improve cardiac resynchronization therapy,” Nat. Rev. Cardiol., vol. 11, no. 8, pp. 481–493, Aug. 2014, doi: 10.1038/nrcardio.2014.67.

[4] C. Daubert, N. Behar, R. P. Martins, P. Mabo, and C. Leclercq, “Avoiding non-responders to cardiac resynchronization therapy: a practical guide,” Eur. Heart J., vol. 38, no. 19, p. ehw270, Jul. 2016, doi: 10.1093/eurheartj/ehw270.

[5] J. Marek, J. Gandalovičová, E. Kejřová, M. Pšenička, A. Linhart, and T. Paleček, “Echocardiography and cardiac resynchronization therapy,” Cor Vasa, vol. 58, no. 3, pp. e340–e351, Jun. 2016, doi: 10.1016/j.crvasa.2015.08.001.

[6] F. Z. Khan et al., “Targeted Left Ventricular Lead Placement to Guide Cardiac Resynchronization Therapy,” J. Am. Coll. Cardiol., vol. 59, no. 17, pp. 1509–1518, Apr. 2012, doi: 10.1016/j.jacc.2011.12.030.

[7] A. Vitarelli, P. Franciosa, and S. Rosanio, “Tissue Doppler Imaging in the assessment of selection and response from cardiac resynchronization therapy,” Eur. J. Echocardiogr., vol. 8, no. 5, pp. 309–316, Oct. 2007, doi: 10.1016/j.euje.2006.12.005.

[8] E. S. Chung et al., “Results of the Predictors of Response to CRT (PROSPECT) Trial,” Circulation, vol. 117, no. 20, pp. 2608–2616, May 2008, doi: 10.1161/CIRCULATIONAHA.107.743120.

[9] F. Leyva, “Cardiac resynchronization therapy guided by cardiovascular magnetic resonance,” J. Cardiovasc. Magn. Reson., vol. 12, no. 1, p. 64, Dec. 2010, doi: 10.1186/1532-429X-12-64.

[10] R. J. Taylor, F. Umar, J. R. Panting, B. Stegemann, and F. Leyva, “Left ventricular lead position, mechanical activation, and myocardial scar in relation to left ventricular reverse remodeling and clinical outcomes after cardiac resynchronization therapy: A feature-tracking and contrast-enhanced cardiovascular magnetic r,” Hear. Rhythm, vol. 13, no. 2, pp. 481–489, Feb. 2016, doi: 10.1016/j.hrthm.2015.10.024.s

[11] E. R. McVeigh, F. W. Prinzen, B. T. Wyman, J. E. Tsitlik, H. R. Halperin, and W. C. Hunter, “Imaging asynchronous mechanical activation of the paced heart with tagged MRI,” Magn. Reson. Med., vol. 39, no. 4, pp. 507–513, Apr. 1998, doi: 10.1002/mrm.1910390402.

[12] D. A. Auger et al., “Imaging left-ventricular mechanical activation in heart failure patients using cine DENSE MRI: Validation and implications for cardiac resynchronization therapy,” J. Magn. Reson. Imaging, vol. 46, no. 3, pp. 887–896, Sep. 2017, doi: 10.1002/jmri.25613.

[13] A. K. Shetty et al., “Cardiac magnetic resonance-derived anatomy, scar, and dyssynchrony fused with fluoroscopy to guide LV lead placement in cardiac resynchronization therapy: a comparison with acute haemodynamic measures and echocardiographic reverse remodelling,” Eur. Hear. J. - Cardiovasc. Imaging, vol. 14, no. 7, pp. 692–699, Jul. 2013, doi: 10.1093/ehjci/jes270.

[14] Q. A. Truong et al., “Utility of dual-source computed tomography in cardiac resynchronization therapy—DIRECT study,” Hear. Rhythm, vol. 15, no. 8, pp. 1206–1213, Aug. 2018, doi: 10.1016/j.hrthm.2018.03.020.

[15] J. M. Behar et al., “Comprehensive use of cardiac computed tomography to guide left ventricular lead placement in cardiac resynchronization therapy,” Hear. Rhythm, vol. 14, no. 9, pp. 1364–1372, Sep. 2017, doi: 10.1016/j.hrthm.2017.04.041.

[16] J. Gould et al., “Feasibility of intraprocedural integration of cardiac CT to guide left ventricular lead implantation for CRT upgrades,” J. Cardiovasc. Electrophysiol., vol. 32, no. 3, pp. 802–812, Mar. 2021, doi: 10.1111/jce.14896.

[17] D. B. Fyenbo et al., “Transmural Myocardial Scar Assessed by Cardiac Computed Tomography,” J. Comput. Assist. Tomogr., vol. 43, no. 2, pp. 312–316, 2019, doi: 10.1097/RCT.0000000000000824.

[18] A. Sommer et al., “Empiric versus imaging guided left ventricular lead placement in cardiac resynchronization therapy (ImagingCRT): study protocol for a randomized controlled trial,” Trials, vol. 14, no. 1, p. 113, 2013, doi: 10.1186/1745-6215-14-113.

[19] A. Sommer et al., “Multimodality imaging-guided left ventricular lead placement in cardiac resynchronization therapy: a randomized controlled trial,” Eur. J. Heart Fail., vol. 18, no. 11, pp. 1365–1374, Nov. 2016, doi: 10.1002/ejhf.530.

[20] G. M. Colvert et al., “Novel 4DCT Method to Measure Regional Left Ventricular Endocardial Shortening Before and After Transcatheter Mitral Valve Implantation,” Struct. Hear., vol. 5, no. 4, pp. 410–419, Jul. 2021, doi: 10.1080/24748706.2021.1934617.

[21] A. Manohar, G. M. Colvert, A. Schluchter, F. Contijoch, and E. R. McVeigh, “Anthropomorphic left ventricular mesh phantom: a framework to investigate the accuracy of SQUEEZ using Coherent Point Drift for the detection of regional wall motion abnormalities,” J. Med. Imaging, vol. 6, no. 04, p. 1, Dec. 2019, doi: 10.1117/1.JMI.6.4.045001.

[22] P. A. Yushkevich et al., “User-guided 3D active contour segmentation of anatomical structures: Significantly improved efficiency and reliability,” Neuroimage, vol. 31, no. 3, pp. 1116–1128, Jul. 2006, doi: 10.1016/j.neuroimage.2006.01.015.

[23] E. R. McVeigh et al., “Regional myocardial strain measurements from 4DCT in patients with normal LV function,” J. Cardiovasc. Comput. Tomogr., vol. 12, no. 5, pp. 372–378, Sep. 2018, doi: 10.1016/j.jcct.2018.05.002.

[24] A. K. Feeny et al., “Machine Learning Prediction of Response to Cardiac Resynchronization Therapy,” Circ. Arrhythmia Electrophysiol., vol. 12, no. 7, pp. 1–12, Jul. 2019, doi: 10.1161/CIRCEP.119.007316.

[25] B. E. Nett, J. D. Pack, and D. Okerlund, “Task based assessment of a motion compensation algorithm via simulation of a moving stenotic vessel,” in Medical Imaging 2013: Physics of Medical Imaging, Mar. 2013, no. 86682, p. 86682B, doi: 10.1117/12.2006593.

[26] A. Manohar, J. D. Pack, A. J. Schluchter, and E. R. McVeigh, “Four-dimensional computed tomography of the left ventricle, Part II: estimation of mechanical activation times,” Med. Phys., Feb. 2022, doi: 10.1002/mp.15550.

[27] M. B. Kronborg et al., “Left ventricular regional remodeling and lead position during cardiac resynchronization therapy,” Hear. Rhythm, vol. 15, no. 10, pp. 1542–1549, Oct. 2018, doi: 10.1016/j.hrthm.2018.04.012.

[28] L. Sugeng et al., “Quantitative assessment of left ventricular size and function: Side-by-side comparison of real-time three-dimensional echocardiography and computed tomography with magnetic resonance reference,” Circulation, vol. 114, no. 7, pp. 654–661, 2006, doi: 10.1161/CIRCULATIONAHA.106.626143.

[29] C. Cortes and V. Vapnik, “Support-vector networks,” Mach. Learn., vol. 20, no. 3, pp. 273–297, Sep. 1995, doi: 10.1007/BF00994018.

[30] E. Bauer and R. Kohavi, “Empirical comparison of voting classification algorithms: bagging, boosting, and variants,” Mach. Learn., vol. 36, no. 1, pp. 105–139, 1999, doi: 10.1023/a:1007515423169.

[31] S. Sukhanov, A. Merentitis, C. Debes, J. Hahn, and A. M. Zoubir, “Bootstrap-based SVM aggregation for class imbalance problems,” in 2015 23rd European Signal Processing Conference (EUSIPCO), Aug. 2015, pp. 165–169, doi: 10.1109/EUSIPCO.2015.7362366.

[32] K. Matsumoto et al., “Reverse remodelling induces progressive ventricular resynchronization after cardiac resynchronization therapy ‘from vicious to virtuous cycle,’” Eur. J. Echocardiogr., vol. 12, no. 10, pp. 782–789, Oct. 2011, doi: 10.1093/ejechocard/jer143.

[33] M. G. St John Sutton et al., “Effect of Cardiac Resynchronization Therapy on Left Ventricular Size and Function in Chronic Heart Failure,” Circulation, vol. 107, no. 15, pp. 1985–1990, Apr. 2003, doi: 10.1161/01.CIR.0000065226.24159.E9.

[34] R. Ramachandran, X. Chen, C. M. Kramer, F. H. Epstein, and K. C. Bilchick, “Singular Value Decomposition Applied to Cardiac Strain from MR Imaging for Selection of Optimal Cardiac Resynchronization Therapy Candidates,” Radiology, vol. 275, no. 2, pp. 413–420, May 2015, doi: 10.1148/radiol.14141578.

[35] A. C. Kydd, F. Z. Khan, D. O’Halloran, P. J. Pugh, M. S. Virdee, and D. P. Dutka, “Radial Strain Delay Based on Segmental Timing and Strain Amplitude Predicts Left Ventricular Reverse Remodeling and Survival After Cardiac Resynchronization Therapy,” Circ. Cardiovasc. Imaging, vol. 6, no. 2, pp. 177–184, Mar. 2013, doi: 10.1161/CIRCIMAGING.112.000191.

[36] C. Chen et al., “Deep Learning for Cardiac Image Segmentation: A Review,” Front. Cardiovasc. Med., vol. 7, no. March, Mar. 2020, doi: 10.3389/fcvm.2020.00025.

[37] D. M. Vigneault, F. Contijoch, C. P. Bridge, K. Lowe, C. Jan, and E. R. McVeigh, “M-SiSSR: Regional Endocardial Function Using Multilabel Simultaneous Subdivision Surface Registration,” in Functional Imaging and Modeling of the Heart, 2021, pp. 242–252, doi: 10.1007/978-3-030-78710-3_24.

[38] C. H. McCollough, A. N. Primak, N. Braun, J. Kofler, L. Yu, and J. Christner, “Strategies for Reducing Radiation Dose in CT,” Radiol. Clin. North Am., vol. 47, no. 1, pp. 27–40, Jan. 2009, doi: 10.1016/j.rcl.2008.10.006.

[39] T. J. Stocker et al., “Reduction in radiation exposure in cardiovascular computed tomography imaging: results from the PROspective multicenter registry on radiaTion dose Estimates of cardiac CT angIOgraphy iN daily practice in 2017 (PROTECTION VI),” Eur. Heart J., vol. 39, no. 41, pp. 3715–3723, Nov. 2018, doi: 10.1093/eurheartj/ehy546.

[40] M. M. Lell and M. Kachelrieß, “Recent and Upcoming Technological Developments in Computed Tomography,” Invest. Radiol., vol. 55, no. 1, pp. 8–19, Jan. 2020, doi: 10.1097/RLI.0000000000000601.

[41] C. Leclercq et al., “Systolic Improvement and Mechanical Resynchronization Does Not Require Electrical Synchrony in the Dilated Failing Heart With Left Bundle-Branch Block,” Circulation, vol. 106, no. 14, pp. 1760–1763, Oct. 2002, doi: 10.1161/01.CIR.0000035037.11968.5C.

[42] R. H. Helm et al., “Cardiac Dyssynchrony Analysis Using Circumferential Versus Longitudinal Strain,” Circulation, vol. 111, no. 21, pp. 2760–2767, May 2005, doi: 10.1161/CIRCULATIONAHA.104.508457.

[43] B. Ambale-Venkatesh et al., “Left ventricular shape predicts different types of cardiovascular events in the general population,” Heart, vol. 103, no. 7, pp. 499–507, Apr. 2017, doi: 10.1136/heartjnl-2016-310052.

[44] B. T. Wyman, W. C. Hunter, F. W. Prinzen, and E. R. McVeigh, “Mapping propagation of mechanical activation in the paced heart with MRI tagging,” Am. J. Physiol. Circ. Physiol., vol. 276, no. 3, pp. H881–H891, Mar. 1999, doi: 10.1152/ajpheart.1999.276.3.H881.

[45] B. T. Wyman, W. C. Hunter, F. W. Prinzen, O. P. Faris, and E. R. McVeigh, “Effects of single-and biventricular pacing on temporal and spatial dynamics of ventricular contraction,” Am. J. Physiol. Circ. Physiol., vol. 282, no. 1, pp. H372–H379, Jan. 2002, doi: 10.1152/ajpheart.2002.282.1.H372.

[46] H. He and Y. Ma, Imbalanced Learning: Foundations, Algorithms, and Applications. Wiley, 2013.

[47] A. R. T. Donders, G. J. M. G. van der Heijden, T. Stijnen, and K. G. M. Moons, “Review: A gentle introduction to imputation of missing values,” J. Clin. Epidemiol., vol. 59, no. 10, pp. 1087–1091, Oct. 2006, doi: 10.1016/j.jclinepi.2006.01.014.

[48] W.-J. Lin and J. J. Chen, “Class-imbalanced classifiers for high-dimensional data,” Brief. Bioinform., vol. 14, no. 1, pp. 13–26, Jan. 2013, doi: 10.1093/bib/bbs006.

