## Supplemental Methods for "Prediction of CRT Response Using a Lead Placement Score Derived from 4DCT"

S1. Global Features of LV Mechanics

*S1.1 LV Volumes and Ejection Fraction*

The LV volume was computed as a function of time across the cardiac cycle for all 20 times frames in each 4D dataset by summing the segmented LV blood pool voxels in each time frame. LVEF was then calculated as:

$$LVEF = \frac{EDV - ESV}{EDV}, \quad (1)$$

where $EDV$ is the LV volume in mL at end diastole and $ESV$ is the LV volume in mL at end systole.

*S1.2 Circumferential Uniformity Ratio Estimate*

An additional feature used in this work is the circumferential uniformity ratio estimate (CURE) [41], [42]. CURE is a metric that describes the synchrony or dyssynchrony of contraction in the circumferential direction of a particular short axis slice of the LV. A spatiotemporal matrix containing $RS_{CT}$ values was constructed, with the columns representing time frames of the cardiac cycle and the rows representing the spatial sampling of $RS_{CT}$ in the circumferential direction of a slice of the LV endocardium. To obtain an estimate of CURE that was less sensitive to noise, a singular value decomposition (SVD) was performed on this matrix to smooth out noise and to identify the primary spatiotemporal pattern of $RS_{CT}$ in the particular LV slice. Only the rank-1 SVD approximation was retained. A modified CURE value based on this rank-1 SVD approximation, called CURE-SVD [34], was calculated as the ratio of the amplitude of the DC component to the sum of the amplitudes of the DC and the first order sinusoidal frequency components, given by the equation:

$$CURE - SVD = \frac{f_0}{f_0 + f_1}, \quad (2)$$

where $f_0$ is the amplitude of the DC component and $f_1$ is the amplitude of the first order sinusoidal component. These frequency components were derived from a Fourier transform analysis of the circumferential distribution of $RS_{CT}$ at a particular time frame of the cardiac cycle (Fourier transform of the signal from a particular column of the rank-1 SVD matrix). The Fourier transform analysis was performed using the built-in *fft* function in MATLAB (MathWorks Inc., Natick, MA; v9.8 – R2020a). The above process of constructing a spatiotemporal matrix of $RS_{CT}$, deriving its rank-1 SVD approximation, and calculating CURE-SVD was repeated for all defined short axis slices of the LV endocardium. The final CURE-SVD value for a subject was computed as the average of all CURE-SVD values across all defined LV short axis slices.

*S1.3 Left Ventricular Sphericity Index*

Left ventricular sphericity index (LVSI) is a measure that captures the degree of sphericity of the left ventricle and was not used in previous 4DCT studies to guide CRT. It is defined as the ratio of EDV to the volume of a sphere whose diameter is equal to the major axis (long axis) of the LV at end diastole [43], given by the formula:

$$LVSI = \frac{EDV}{\frac{\pi * L^3}{6}}, \quad (3)$$

where $L$ is the length of the long axis of the LV at end diastole.

S2. Regional Features of LV Mechanics

*S2.1 Regional Endocardial Shortening*

Regional endocardial shortening ($RS_{CT}$) from 4DCT images [20], [23] was measured according to the following formula:

$$RS_{CT}(\nu, t) = \sqrt{\frac{A(\nu,t)}{A(\nu,t=0)}} - 1, \quad (4)$$

where $A(\nu, t)$ is the area of an endocardial patch $\nu$ at time $t$ of the cardiac cycle and $A(\nu, t = 0)$ is the area of the same endocardial patch $\nu$ at end diastole ($t = 0$). The location of each patch $\nu$ was tracked over the heart cycle using a point set registration algorithm [21]; the average patch size at end diastole was 2.6 ± 0.5 $mm^2$. For each time frame of the cardiac cycle, the $RS_{CT}$ values were mapped onto the endocardial surface, yielding a high-resolution estimate of regional endocardial shortening.

For each subject, three features derived from the spatiotemporal distribution of $RS_{CT}$ were defined (also see Fig. S1):

1. Peak regional shortening ($PRS_{CT}$): maximum shortening achieved in a spatial segment of the LV. This feature correlates strongly with the existence of myocardial infarction.
2. Time to peak regional shortening ($TPRS_{CT}$): normalized time of the cardiac cycle at which peak shortening occurs in a spatial segment of the LV.
3. Maximum pre-stretch of regional shortening ($MSRS_{CT}$): maximum endocardial pre-stretch ($RS_{CT} > 0$) in a spatial segment of the LV. If pre-stretch does not occur, then $MSRS_{CT}$ is 0. This feature correlates strongly with late mechanical activation, and poor myocardial function in that segment.

*S2.2 Time to Onset of Shortening*

The "time to onset of shortening" (TOS) is the time at which the endocardium *begins* to contract during systole. It has been shown to be an excellent descriptor of dyssynchrony[11], [44], [45] and a strongly predictive feature for CRT response [12]; this feature was not included in previous studies using 4DCT to guide CRT. There is no universally accepted definition nor method of computation of TOS; for our study, the TOS of a spatial segment of the LV was defined as the time at which the $RS_{CT}$ vs time curve for that segment reached 10% of its dynamic range during systole. The dynamic range of shortening during systole was defined as the range from maximum pre-stretch ($MSRS_{CT}$) to peak shortening ($PRS_{CT}$); if no pre-stretch occurred, it was defined as the range of shortening from end diastole ($RS_{CT} = 0$) to peak shortening ($RS_{CT} = PRS_{CT}$). For LV spatial segments with very low function, the TOS was not estimated due to insufficient dynamic range. Figure S1 illustrates the values of $PRS_{CT}$, $TPRS_{CT}$, $MSRS_{CT}$, and TOS for an exemplar $RS_{CT}$ vs time curve from our 4DCT studies. Curves like this were measured over 90 contiguous local regions of the LV; each region represents approximately 2 $cm^2$ of endocardial surface for a normal ventricle.

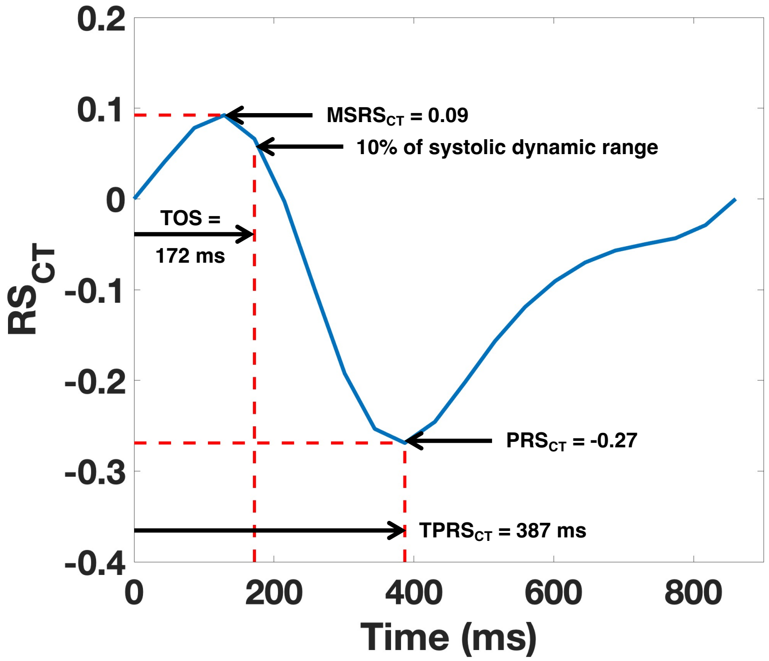


**Fig. S1. Measurements of maximum pre-stretch of regional shortening ($MSRS_{CT}$), time to onset of shortening (TOS), peak regional shortening ($PRS_{CT}$), and time to peak regional shortening ($TPRS_{CT}$) for an exemplar $RS_{CT}$ vs time curve for a single location on the LV.** $RS_{CT}$: regional endocardial shortening.

S3. Support Vector Machine Training and Lead Placement Score

The built-in *fitcsvm* function in MATLAB (Statistics and Machine Learning Toolbox, MathWorks Inc., Natick, MA; v9.8 – R2020a) was used to train the SVM model (‘kernel’: linear; ‘Standardize’: true; ‘Prior’: uniform; all other parameters were set to default). The prior probability input parameter was set to ‘uniform’ to handle the class imbalance between responders (n=52) and non-responders (n=30) in our dataset; thus, proportionately weighting the cost of misclassification [46]. A bagging [30] approach with 1000 iterations was implemented for training; for each iteration, an SVM model was trained on a bootstrapped dataset that was sampled with replacement. Each bootstrapped dataset had the same number of subjects as the original dataset and the trained SVM model obtained was a vector of 12 feature coefficients. Nine TOS-right values were indeterminable and were imputed via the method of mean imputation [47] for training. The final model was derived by averaging the feature coefficients of the individually trained models on the 1000 bootstrapped datasets (the mean values of the feature coefficients converged around the 350th iteration).

The LPS was calculated for all subjects in the study according to the formula:

$$LPS_i = \boldsymbol{\beta} \cdot \boldsymbol{x_i} + b, \tag{5}$$

where $\boldsymbol{\beta}$ is the vector of mean feature coefficients, $\boldsymbol{x}$ is the feature vector values at the location of the left and the right leads, $b$ is the bias term, and $i = 1,2,3, \dots, N$ is an index for the $N$ subjects used in the study. The LPS value that maximized the geometric mean (g-mean) was chosen as the "optimal" threshold for the prediction of responders and non-responders. The g-mean is a performance metric that balances the correct prediction of both the majority (CRT responders) and the minority (CRT non-responders) classes [48]; it is defined as the square root of the product of the sensitivity and the specificity.
